# A data-driven metapopulation model for the Belgian COVID-19 epidemic: assessing the impact of lockdown and exit strategies

**DOI:** 10.1101/2020.07.20.20157933

**Authors:** Pietro Coletti, Pieter Libin, Oana Petrof, Lander Willem, Steven Abrams, Sereina A. Herzog, Christel Faes, James Wambua, Elise Kuylen, COVID-19 team the SIMID, Philippe Beutels, Niel Hens

**Affiliations:** Data Science Institute, I-BioStat, UHasselt, Hasselt, Belgium; Global Health Institute, Department of Epidemiology and Social Medicine, University of Antwerp, Antwerp, Belgium; Centre for Health Economic Research and Modelling Infectious Diseases, Vaccine & Infectious Disease Institute, University of Antwerp, Antwerp, Belgium; The members of the SIMID-COVID-19 team will be listed in the peer-reviewed paper; School of Public Health and Community Medicine, The University of New South Wales, Sydney, Australia

## Abstract

**Background:** In response to the ongoing COVID-19 pandemic, several countries adopted measures of social distancing to a different degree. For many countries, after successfully curbing the initial wave, lockdown measures were gradually lifted. In Belgium, such relief started on May 4th with phase 1, followed by several subsequent phases over the next few weeks.

**Methods:** We analysed the expected impact of relaxing stringent lockdown measures taken according to the phased Belgian exit strategy. We developed a stochastic, data-informed, meta-population model that accounts for mixing and mobility of the age-structured population of Belgium. The model is calibrated to daily hospitalization data and serological data and is able to reproduce the outbreak at the national level. We consider different scenarios for relieving the lockdown, quantified in terms of relative reductions in pre-pandemic social mixing and mobility. We validate our assumptions by making comparisons with social contact data collected during and after the lockdown.

**Results:** Our model is able to successfully describe the initial wave of COVID-19 in Belgium and identifies interactions during leisure/other activities as pivotal in the exit strategy. Indeed, we find a smaller impact of school re-openings as compared to restarting leisure activities and re-openings of work places. We also assess the impact of case isolation of new (suspected) infections, and find that it allows re-establishing relatively more social interactions while still ensuring epidemic control. Scenarios predicting a second wave of hospitalizations were not observed, suggesting that the per-contact probability of infection has changed with respect to the pre-lockdown period.

**Conclusions:** Community contacts are found to be most influential, followed by professional contacts and school contacts, respectively, for an impending second wave of COVID-19. Regular re-assessment is crucial to adjust to evolving behavioral changes that can affect epidemic diffusion. In addition to social distancing, sufficient capacity for extensive testing and contact tracing is essential for successful mitigation.

## 1 Introduction

The COVID-19 pandemic has put a massive burden on modern society. While the global death toll of the virus has risen above 500,000 reported deaths on the 15th of July [1], several countries are evaluating strategies to cope with the virus on the medium to long term. As neither a vaccine nor adequate therapeutic options are available at this time, non-pharmaceutical interventions have been proven effective in reducing the pressure on healthcare systems [2, 3, 4, 5, 6]. After a massive implementation of lockdown measures, affecting as much as one third of the global world population [7], governments have eased some of the social distancing measures. After imposing a partial lockdown on March 14th and a full lockdown on March 18th, the Belgian government curtailed some of these measures with a plan for a gradual reopening over several weeks, starting from the 4th of May. The absence of substantial population immunity after this first wave of COVID-19 in Belgium [8] increases the risk of subsequent large-scale outbreaks when interventions are relaxed which could result, when not contained, in new COVID-19 waves with large numbers of new confirmed and hospitalized persons. In this context, data-driven models of disease spread can provide useful insights into the expected impact of easing non-pharmaceutical interventions [6, 9, 2]. Here we revisit the different scenarios which could have unfolded as a consequence of easing lockdown measures based on a data-driven metapopulation model for Belgium for COVID-19 [10] and compare the expected and realized epidemic trajectories. At the same time, we validate the modelled scenarios with social contact data collected during and after lockdown.

## 2 Methodology

We constructed a meta-population model for COVID-19, in order to study the Belgian epidemic. The model reproduces the demography of *children* (0-18 years) and *adults* (19 years and above) in the different Belgian municipalities [11]. Publicly available data [12, 13] from a social contact survey conducted in Flanders (Belgium) anno 2010-2011 is used to inform mixing patterns of the population [14, 15, 16]. Mobility data retrieved from the Belgian census [17] is used to reconstruct mobility fluxes due to school attendance and work. A stochastic compartmental model is used to describe the spread of COVID-19 in the population within each patch of the system. The model is fitted to national hospitalization data [18] and serial serological survey data [8].

### 2.1 Compartmental patch model

We use an extended *SEIR* stochastic compartmental model (Figure 1) in which we distinguish pre-symptomatic (*I*_*p*_), asymptomatic (*I*_*a*_), and symptomatic (*I*_ms_ and *I*_ss_) transmission by assuming different transmission rates, governed by different contact patterns. In particular, we assume that symptomatic individuals (both *mildly* symptomatic *I*_ms_ and *severely* symptomatic *I*_ss_) reduce their number of contacts (following observations made during the 2009 Influenza pandemic [19]) and their commute (school/work) mobility. A fraction of symptomatic adults can show severe symptoms and therefore require hospitalization (*H*) [5, 20]. Once this happens, we assume that they cannot further infect other people due to isolation measures [21].

**Figure 1:**
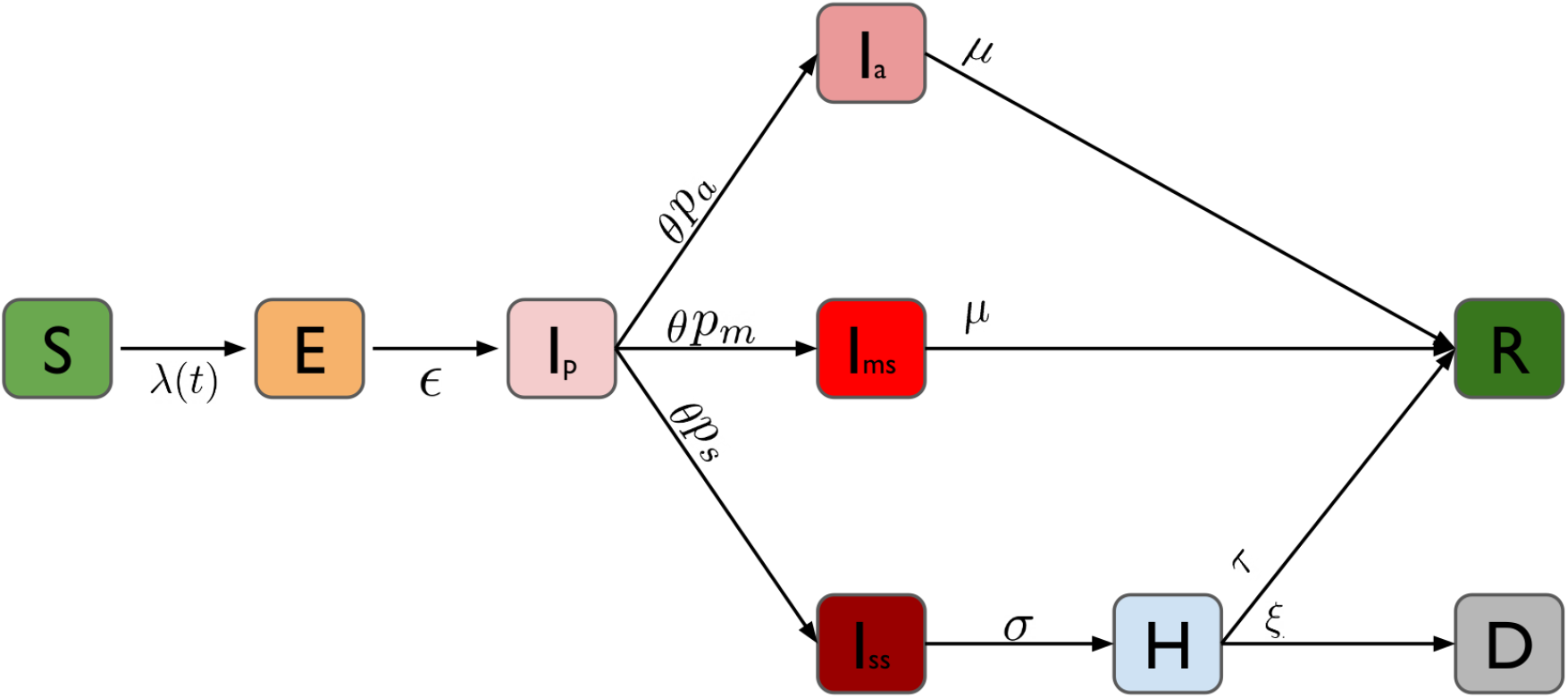
Schematic representation of the compartmental model: Individuals start as susceptible (*S*) and can become exposed to the disease (*E*) when interacting with infected individuals (*I*_*p*_, *I*_*a*_, *I*_ms_ and *I*_ss_). After a latent period, exposed individuals enter a pre-symptomatic phase (*I*_*p*_), after which they can either become symptomatic (*I*_ms_ and *I*_ss_) or remain without symptoms (*I*_*a*_). Symptomatic individuals can develop *mild* symptoms (*I*_ms_) or severe symptoms (*I*_ss_). When symptoms are severe, they are hospitalized (*H*). The final outcome of infected individuals is either recovery (*R*) or death (*D*).

We assume that children have a 50% lower susceptibility to infection compared to adults [4, 22, 23]. Table 1 shows a summary of the model parameters and the distributional assumptions thereabout.

**Table 1:** Overview of the model parameters.

| Quantity | Median (95% CI) | Distribution | Source |
| --- | --- | --- | --- |
| Latent period ($\epsilon$) | 1.4 days ([0:7] days) | Exponential | [24, 20] |
| Pre-symptomatic period ($\theta$) | 2.4 days ([0:13] days) | Exponential | [24, 20] |
| Children infectivity (wrt adults) | 0.5 | — | [4, 25, 22, 26] |
| Proportion asymptomatic ($p_a$) | 0.5 | — | [25, 26, 27, 28] |
| Proportion mild symptoms ($p_m$) | 0.5/0.425 (children/adults) | — | [25, 22, 26, 27] |
| Proportion severe symptoms ($p_s$) | 0 / 0.0275 (children/adults) | — | [22, 26] |
| Symptomatic/asymptomatic period ($\mu$) | 2.4 days ([0:13] days) | Exponential | [29] |
| Symptom onset to hospitalization ($\sigma$) | 4.7 days ([0:17] days) | Weibull | [29] |
| Hospital admission to death ($\xi$) | 4 days ([1:9] days) | Log-logistic | [29] |
| Hospital admission to recovery ($\tau$) | 5 days ([1:10] days) | Weibull | [29] |

### 2.2 Population mixing

Population mixing is informed by social contact data for different locations (work, home, school, transportation, leisure activity and other) during weekdays and weekends [14, 15], accessible through the Socrates tool [12, 13].

An asymptomatic individual interacts according to a contact matrix that is the sum of the contact matrices that correspond to different locations:

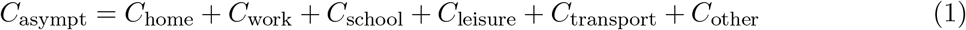

Given the strong age-specific severity of COVID-19, we assume that, when symptomatic, adults reduce their contacts in a location-specific fashion, as reported during the 2009 H1N1 pandemic [19]:

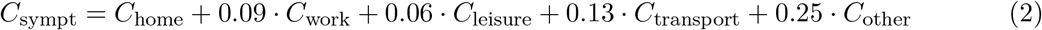

We assume that children do not change behavior when symptomatic, as they are more likely to present fewer and milder symptoms as compared to adults [25, 30, 31]. When intervention measures are implemented (see section 2.4), location-specific contacts are reduced. This has an impact on both *C*_sympt_ and *C*_asympt_, implicitly assuming that a reduction in contacts because of symptoms is the same during the pre-pandemic and intervention period. The contact matrices then become:

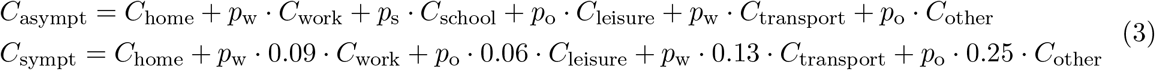

where *p*_w_, *p*_s_, *p*_o_ are the percentages of contacts at work, at school and during leisure/other activities.

### 2.3 Population mobility

Commuting data from the Belgian census [17] is used to infer the daily commuting network among different Belgian municipalities. We assume that intervention measures targeted at reducing contacts at work and at school reduce the mobility of the corresponding age class.

### 2.4 Interventions taken on the 14th of March

Starting from Friday 13th of March at midnight, Belgian authorities have declared the nation-wide closure of schools and universities, together with restaurants, cafes and gyms. Also, public gatherings were banished. On the 17th of March, further dispositions were put in place, limiting mobility of people in addition to closing companies and shops offering non-essential services. We model interventions by reducing mixing and mobility in the population (see section 2.2 and 2.3), with a compliance that increases linearly with time and reaches full compliance on the 23rd of March.

### 2.5 Calibration

To calibrate our model, we used national data on daily hospital admissions [18] and results from serological survey collections [8] to estimate the per-contact transmission probability, the probability of severe symptoms and the reduction of the contact matrix during intervention with respect to the pre-pandemic period. More details on the calibration procedure, including a comparison with the observed seroprevalence data can be found in the Supporting Information (SI).

#### Exit strategies

The Belgian government lifted the lockdown gradually from the 4th of May onward. Table 2 shows a simplified summary of the different phases and their implementation. Changes with respect to the previous phase (i.e. the previous row) are shown in bold. In our scenario analysis we considered three phases:

**Table 2:**
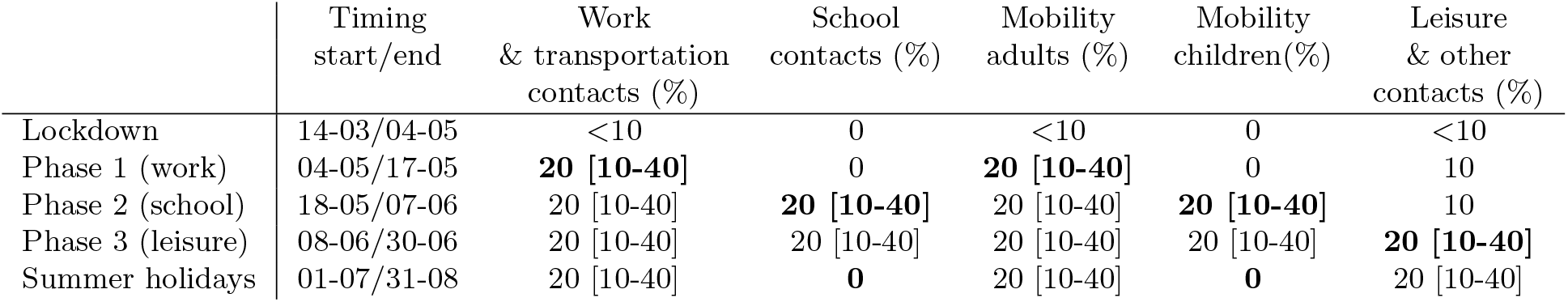
Timing and concepts of lockdown relief. Each phase is implemented incrementally with respect to the previous ones. Bold values highlight the changes with respect to the previous phase (i.e. the previous row). Intermediate parameter values are reported, with the full range between squared brackets.

- Phase 1: from the 4th of May, increasing the contacts made at work and during commuting by adults, to account for the increase of people going back to work. Mobility of adults increases accordingly.
- Phase 2: from the 18th of May, increasing contacts made at school and during commuting by children to account for school re-opening. Mobility of children increases accordingly.
- Phase 3: from the 8th of June, increasing contacts made during leisure and in other locations, to assess the impact of a possible re-opening of leisure activities.

For these phases we considered a compliance that increases linearly with time and reaches full compliance after one week.

#### Case isolation

When extensive contact tracing and testing is available, a viable option for disease mitigation is to isolate individuals that are infected. We assume that, since all contacts of infected individuals are tested, case isolation affects also asymptomatic individuals that would otherwise slip out of usual case-reporting surveillance methods. We present our results in terms of a synthetic quantity, the parameter *α*, that is the percentage of individuals entering the symptomatic or asymptomatic class (*I*_*a*_, *I*_ms_ and *I*_ss_) that are effectively isolated. We assume that these quarantined individuals reduce their contacts by a factor of ten. We do not cover here how to link the target *α* to an optimal strategy for contact tracing and testing that takes into account feasibility thereof in terms of the number of index cases that can be traced, test features (e.g. sensitivity, specificity) that vary as a function of the time since symptom onset, and willingness to report contacts upon being identified as a COVID-19 case [21, 32, 33]. We also assume that no isolation of pre-symptomatic people is implemented (*I*_*p*_). We considered that case isolation can start at the beginning of phase 2 (i.e. on the 18th of May) or at the beginning of phase 3 (i.e. on the 8th of June), to assess the impact of delay in implementation.

## 3 Results

Figure 2(a) shows the daily number of new hospitalizations in the initial phase of the epidemic, compared with our best model fit (red solid line). Hospitalization data up to the 21st of March are consistent with an exponential growth model with a doubling time of 3.09 days (95% CI [3.05 : 3.11]). Combined with our estimated model parameters, this results in a basic reproduction number *R*_0_ = 3.40 (95% CI [3.36 : 3.44]). A strong, periodic effect on the reported number of hospital admissions can be observed, most likely due to delays in hospitalisation during weekends. The no-intervention model is in line with hospitalization data up to the 21th of March, showing that interventions took about one week to impact hospitalizations. Next, we fit to the data up to the 1st of May, as this is required to calibrate the intervention effects (Figure 2(b)). The model including the effect of interventions is compatible with an overall reduction in the total number of contacts of 85% with respect to the period prior the COVID-19 pandemic (see SI for additional information on contact matrices).

**Figure 2:**
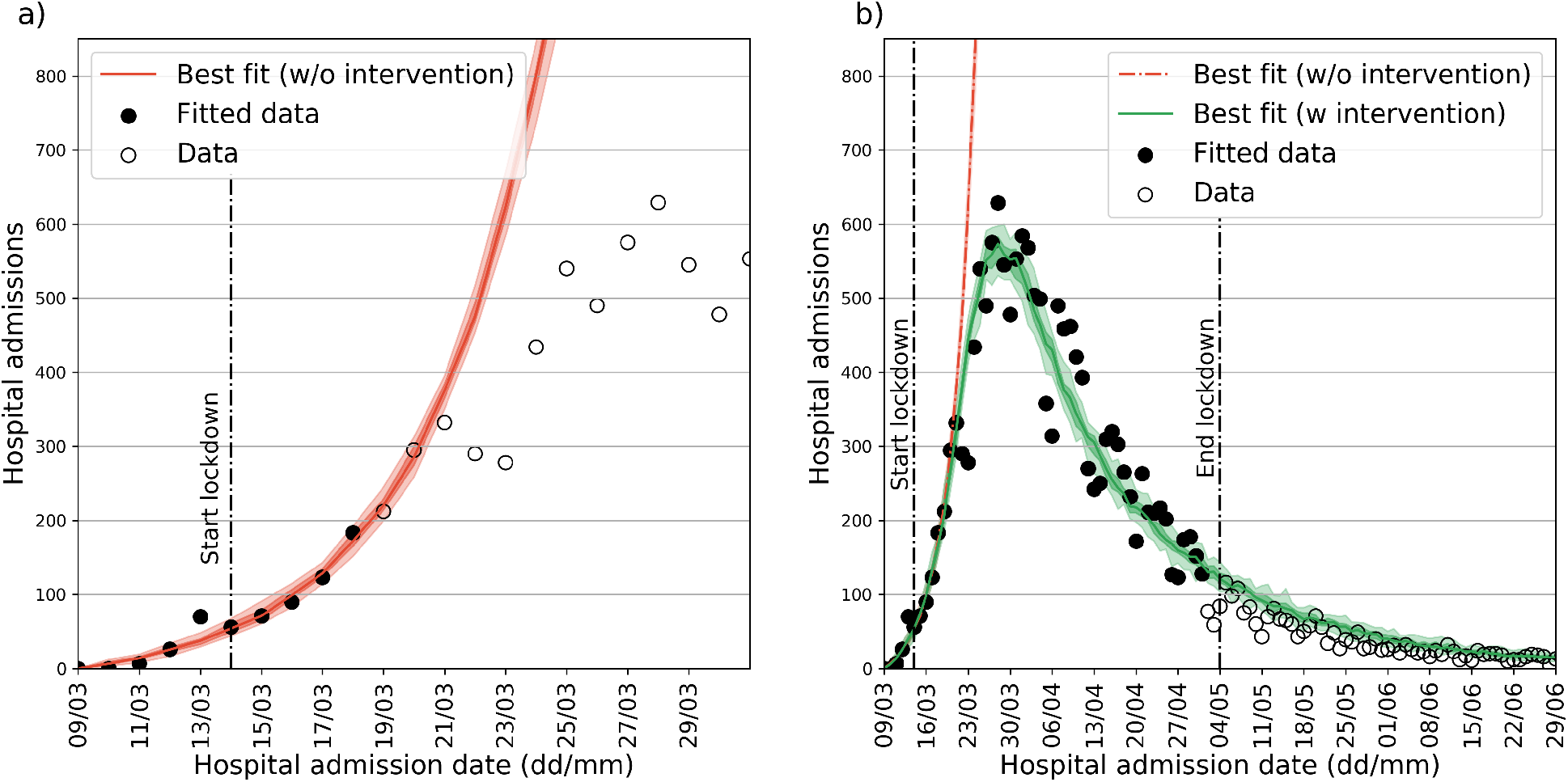
Parameter estimation and model fitting. (**a**): calibration of the pre-intervention phase. Data on hospital admissions is shown in comparison with the best-fit model. Black points are used to calibrate the model in the pre-intervention phase. (**b**): calibration for the intervention phase. Data on hospital admissions is shown in comparison with the best-fit model. Black points are used to calibrate the model in the lockdown phase. In both panels median curves are shown along with 50% confidence intervals (CIs; dark shade) and 95% CI (light shade).

Figure 3 shows the impact of the different phases of the exit strategy, considering different implementations (i.e. parameter values) for each phase, together with a scenario in which lockdown goes on indefinitely. We consider results up to the 31st of August, as considering a longer timeframe would require additional assumptions with regard to social distancing after the summer period. Results for the whole year are reported in the SI (Figure SI 4). In Figure 3(a), at the beginning of phase 1 (4th of May), contacts at work and on transportation are increased, ranging from 10% to 40% of pre-pandemic values. As expected, there is a delay between the implementation of the first phase and its effect on the number of hospital admissions: after 3 weeks the number of hospital admissions stops to decrease as compared to the lockdown scenario. One further week is required to see differences between the three implementations of phase 1. In Figure 3(b) we show the impact of phase 2 (school re-opening) once phase 1 is implemented for the smallest value of contacts at work/transportation considered (10%). The percentage of school contacts ranges from 10% to 40%. In this case, the different curves start to diverge 4 weeks after the re-opening of schools. Summer school holidays, starting on the 1st of July have a considerable (delayed) effect on the number of hospital admissions only in the 40% school contacts scenario. In Figure 3(c) we show the impact of phase 3, once phase 1 and 2 are implemented with the smallest values of the considered parameters. Different implementations of the phase 3 give different results after three weeks. Comparing the three panels, it is clear that changing the implementation of phase 3 has a larger impact than changing implementation of phase 1 or 2. The larger impact of phase 3 (leisure/other activities) is confirmed when comparing all the scenarios we considered. Figure 4 shows the number of daily hospitalization and the cumulative number of hospitalizations up to the 31st of August (results up to the 31st of December are available in SI, Figure SI 4). Results are shown with respect to the scenario with the smallest number of hospitalizations. A smaller impact for school re-opening with respect to work and leisure re-opening is observed, both for peak hospitalizations as for total hospitalizations. Increasing the contacts at school by 10% (i.e. considering a different symbol marker but same color along the y-axis) has a smaller impact than increasing contacts at work (i.e. same symbol, different color along the y-axis) or leisure/other contacts (i.e. moving along the x-axis) of the same amount. Increasing contacts at work has a smaller impact in terms of peak hospitalizations than increasing leisure/other contacts; a similar impact is instead observed for the total number of hospitalizations. When considering results over the whole year (Figure SI 4) the relative increase in the epidemic peak is weakly affected. The total final size, instead, increases for all scenarios, as the daily number of hospitalizations is summed up over a longer period of time, with a larger impact on scenarios with the lowest parameter configurations. However, hospitalization data is compatible with the lockdown scenario (Figure 2) up to the end of June. Comparison of the contact matrices used in the model with the preliminary results of a recent social contact survey [34] targeting Belgian adults during and after the lockdown provides a means to interpret this. Figure 5 shows the measured contact matrices in comparison to one of the simulated scenarios (10% work/transportation contacts, 40% school contacts and 20% of leisure/other contacts). Our model uses a higher number of contacts during phase 1, whereas phase 2 and 3 show general agreement between the model and the empirical data. Since the predicted resurgence in the number of hospitalizations is not observed, this suggests that the proportionality factor between conversational contacts and transmission rates postulated in the so-called social contact hypothesis [35] has changed from the lockdown to the post-lockdown period. This is likely due to behavioral changes (increased hygiene, prominence of outdoor over indoor community contacts, face-mask wearing, etc.) reducing the per-average contact transmission probability. For instance, surveys [36, 34] in Belgium have documented a marked increase in outdoor contacts and face-mask wearing during the three phases of lockdown relief (Figure SI 3), supporting this hypothesis.

**Figure 3:**
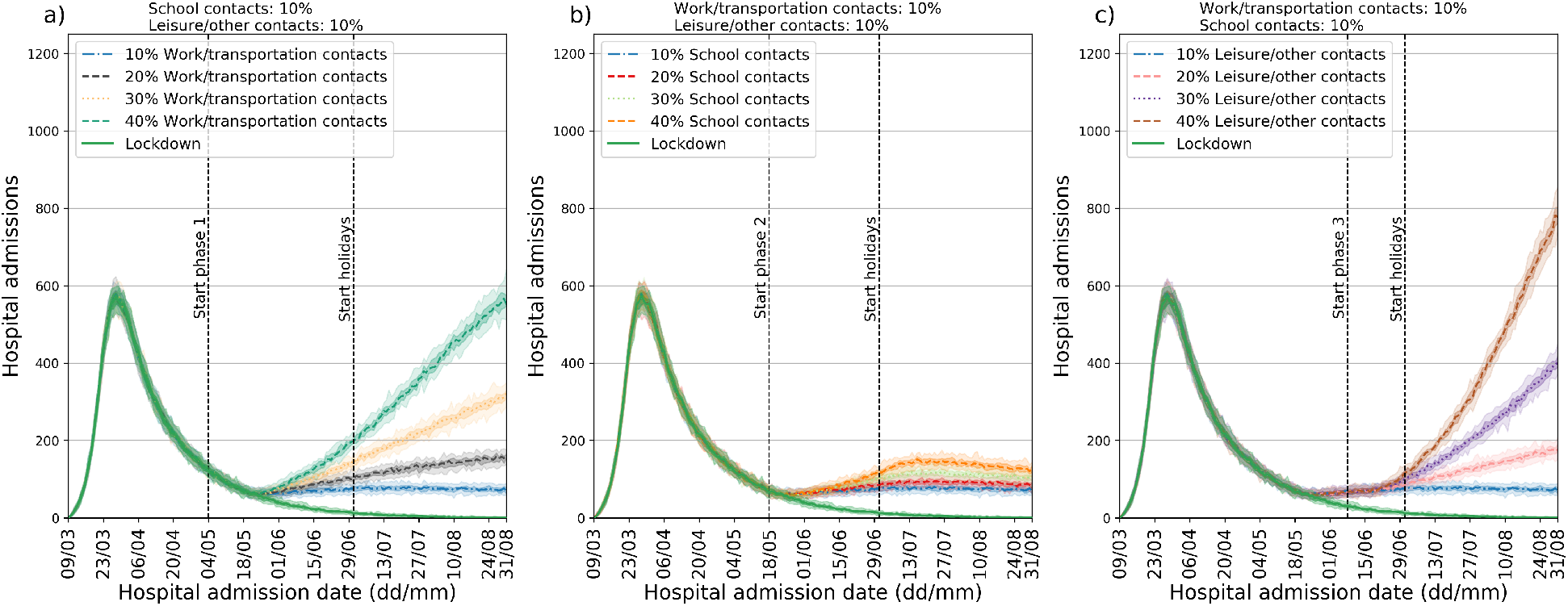
Exit scenarios using different timings and location-specific reductions. (**a**): different implementations of phase 1 (work re-opening). (**b**): different implementations of phase 2 (school re-opening). (**c**): different implementations of phase 3 (leisure re-opening). The top of each panel shows the parameter values used. In all panels median curves are shown along with 50% confidence intervals (dark shade) and 95% CI (light shade). Color-code is consistent across panels, with the same color marking the same scenario in different panels.

**Figure 4:**
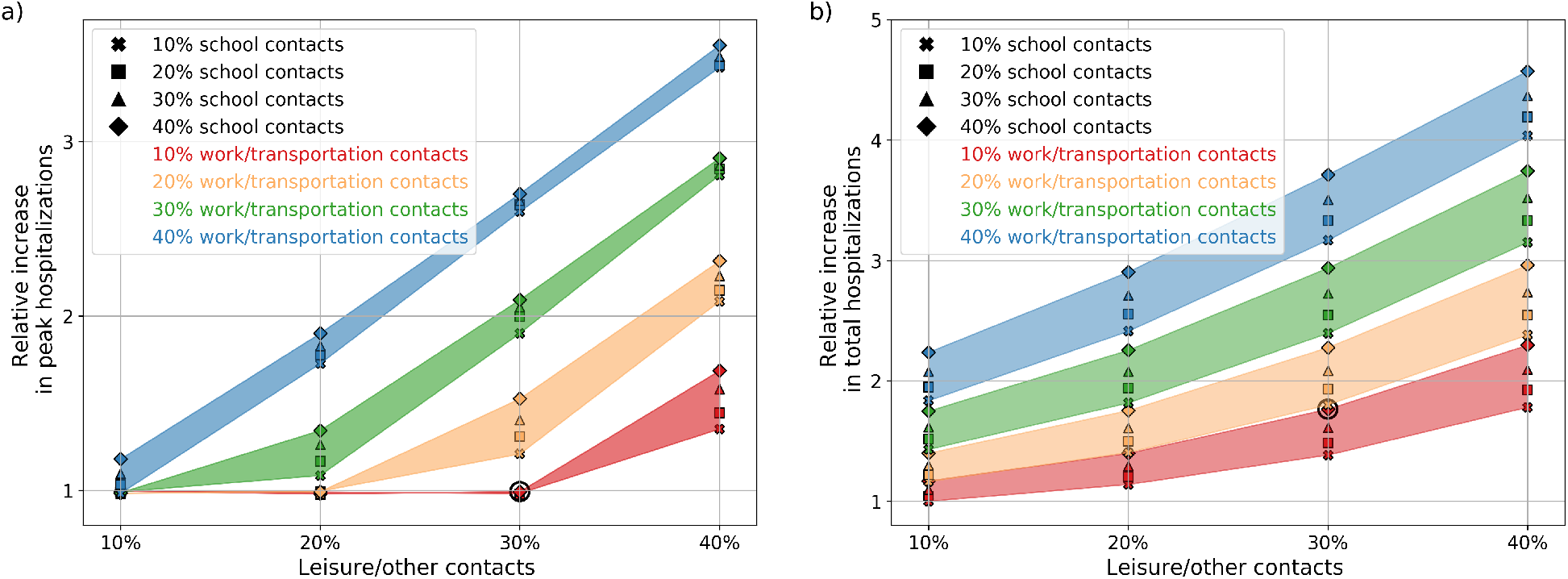
Summary of exit scenarios. (**a**): peak value of daily hospital admissions up to the 31st of August. (**b**): number of hospitalizations up to the 31st of August. In both panels the y-axis shows the relative variation with respect to the best-case (least contacts) scenario. A circle denotes the scenario used in the contact isolation analysis (Figure 6).

**Figure 5:**
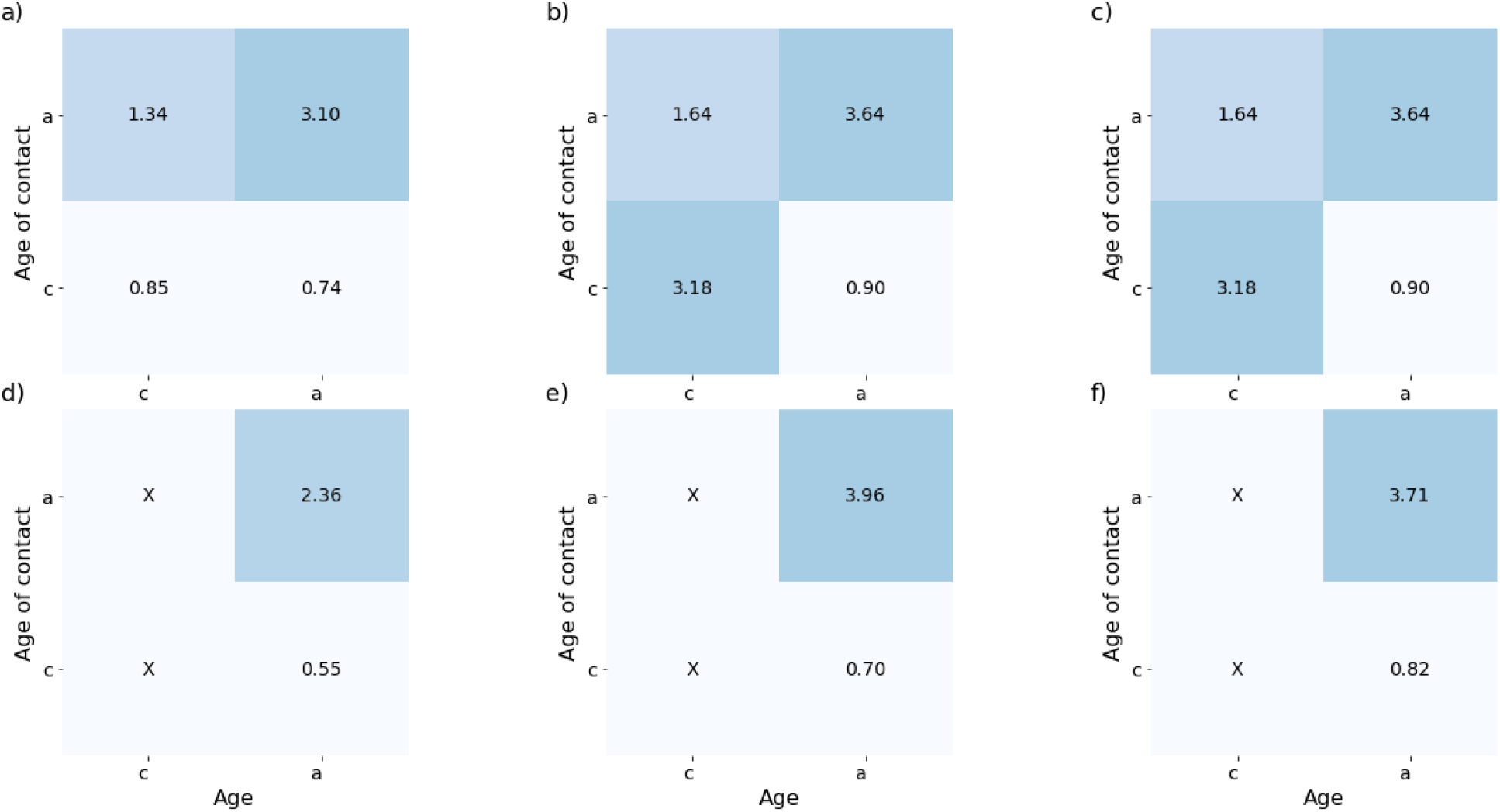
Comparison of model contact matrix and measured ones. (a)-(c): Contact matrices for phase 1 (a), phase 2 (b) and phase 3 (c) in a specific simulated scenario ((10% work/transportation contacts, 40% school contacts and 20% of leisure/other contacts). (d)-(f): Contact matrices for phase 1 (d), phase 2 (e) and phase 3 (f) measured in a survey representative of the Belgian adult population. Contacts of children participants, not measured in the survey, are marked with “X”.

Figure 6 shows the impact of case isolation on the scenario marked with a circle in Figure 4 (10% contacts at work/transportation, 40% contacts at school and 30% leisure/other contacts scenario marked with a circle in Figure 4). The ability to isolate newly infected individuals has a considerable impact on the number of hospital admissions. The isolation of 25% of new cases is able to reduce the expected number of hospital admission at the end of August by 25%. The isolation of twice as many cases (50% instead of 25%) would lead to a reduction of 37% of admissions. Starting case isolation 3 weeks after (at the start of phase 3 instead of phase 2) lessens the reduction to 21% from 25%. A stronger effect of this delay is measured in the 50% case isolation scenario: in this case, starting the isolation at the start of phase 3 decreases the reduction in admissions from 37% to 28%.

**Figure 6:**
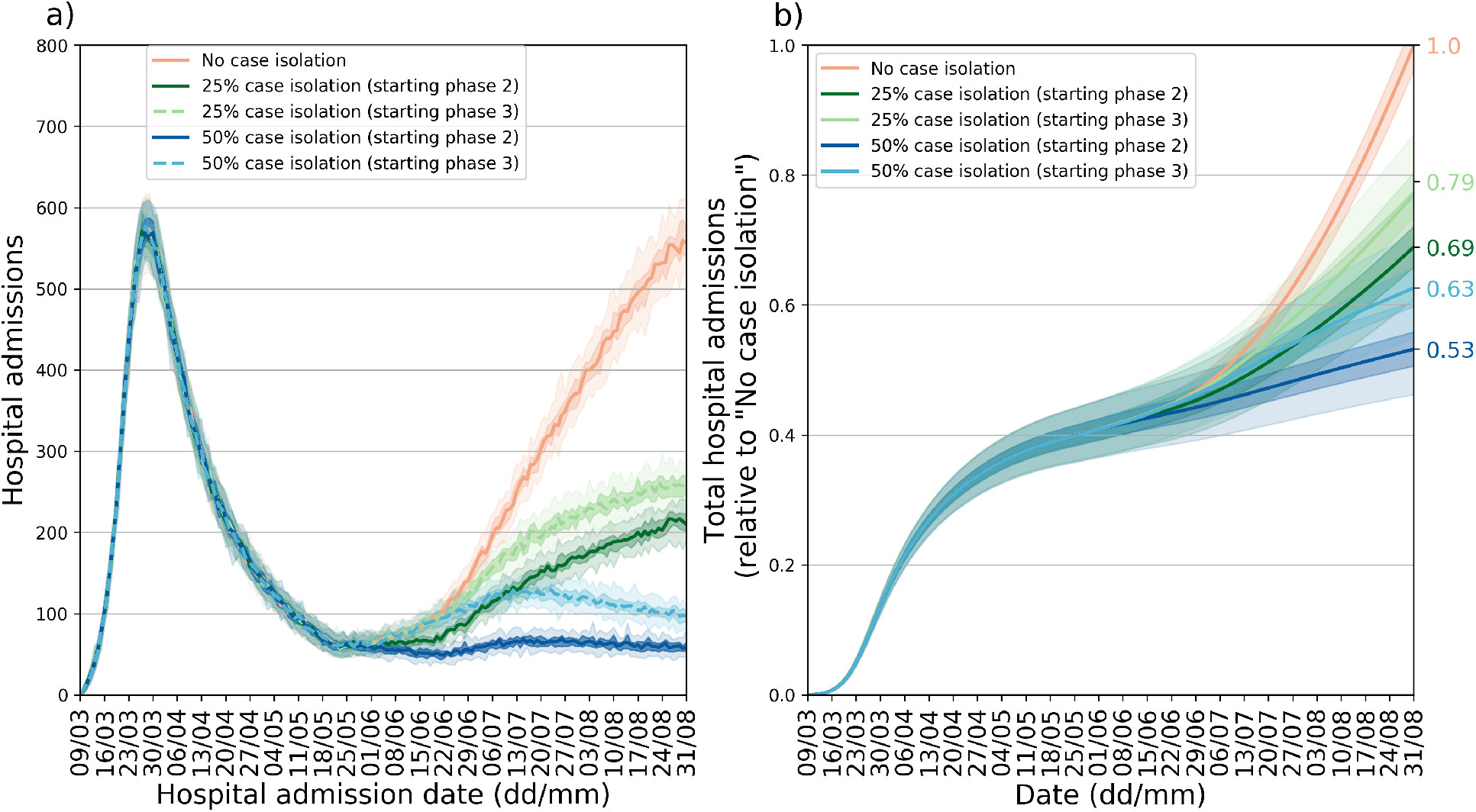
Effect of case isolation in a specific scenario. (**a**): new hospitalizations per day. (**b**): cumulative number of hospitalizations relative to the no case isolation scenario. All curves are obtained considering 40% of working contacts, 40% of contacts at school and 40% of leisure/other contacts with respect to pre-pandemic period (scenario denoted by a black circle in Figure 4). In both panels median curves are shown along with 50% confidence intervals (dark shade) and 95% CI (light shade).

## Discussion

We used a stochastic, discrete time, data-driven meta-population model to predict the impact of lifting the lockdown in three phases. The model includes data on pre-pandemic mobility and mixing, and is calibrated on hospital admissions and seroprevalence data. The initial phase of the COVID-19 epidemic in Belgium is characterized by a fast spread of the disease, with a doubling time of 3.09 days (95% CI [3.05 : 3.14]), in line with values from other countries [5, 2, 37, 38, 39]. Combined with our parameter choices, this results in *R*_0_ = 3.40 (95% CI [3.36 : 3.44]), which lies within the interval estimated in recent meta-analysis (mean = 2.6, standard deviation = 0.54 [40] and mean = 3.28 [39]). Our model appropriately describes hospital admissions during the lockdown period if a strong reduction (85%) in the number of contacts is established. In this situation the number of hospital admissions starts to decrease 3 weeks after the start of the lockdown allowing the healthcare system to cope with ICU demands. At the end of the lockdown, the reproduction number is estimated to be 0.73 (95% CI [0.70 : 0.76]). Such a strong reduction in the average number of contacts marks the disruption that a lockdown has on everyday life. Studies in Wuhan and Shanghai [4] found an even stronger reduction in the number of contacts during lockdown, while a recent survey in the UK [40] measured a reduction of 75%. Preliminary analysis of social contact data collected in Belgium after the lockdown [34] shows similar results as compared to [4] and [40], in line with our modelling results. Adherence to country-specific contact data is paramount, as intervention measures can vary substantially between countries, both in terms of implementation and in terms of compliance. Collecting country specific contact data during the different stages of the epidemic (i.e. before, during and after intervention) is therefore of crucial importance to adequately assess the impact of social distancing [34]. Nevertheless, our knowledge of contact patterns before the COVID-19 crisis can be used to identify the relative impact of introducing social distancing in different locations. In the current analysis this approach was taken, whilst considering a plausible range of reductions in social contacts in different circumstances. According to our model, leisure activities have the largest potential impact on the epidemic profile. This is consistent with leisure/other contacts accounting for 25% to 40% of the total contacts people make, according to representative surveys [41, 42]. However, the absence of a resurgence of hospitalizations by the end of June suggests that there is a smaller per-contact probability of transmission after lockdown with respect to pre-lockdown. This could be due to behavioral changes in how contacts are established (i.e. increased inter-personal distance or the wearing of face masks [43]) after the lockdown or to environmental factors (e.g. humidity and temperature [44]) that could affect transmission. In the light of that, our result are useful in establishing a hierarchy of location-specific contacts, but a careful interpretation of the absolute number of infections is necessary.

We observed less impact of school closure on hospital admissions in contrast to social mixing at work and during transport or leisure activities. First, as expected, school closure leads to observable effects only in those scenarios in which a consistent fraction (i.e. 40% or more) of school contacts are established in the population. Second, as children have a much lower probability of being symptomatic (and as such of being hospitalized) with respect to adults [39], increased diffusion among children increases the observed hospital admissions mostly indirectly, through the increase of infected adults. We tested, as a sensitivity analysis, a scenario in which children have the same susceptibility to the disease: in this case school closure would have a larger impact on the number of infections, especially in the children’s age class. The role of children is still unclear and, although their secondary attack rate in household is similar to the one of adults [30], there is evidence that they present smaller viral load [45, 46, 47, 48] and reduced transmissibility [25, 49], together with a lower number of confirmed cases with respect to adults [22]. This increased susceptibility scenario is therefore unlikely, given the information on COVID-19 we have so far.

In our results, isolation of newly infected individuals has an important impact on epidemic mitigation. Implementing case isolation would allow to re-establish social interactions while still ensuring epidemic containment.We stress here that although we quantified the reduction of spreading potential in terms of number of contacts, this may also come as a combination of different effects, for example when antivirals to be used in the early phase of the infection will become available [50]. Also, a fast setup is crucial: a 3 weeks delay in implementing case isolation leads to a considerable impact on the number of new hospital admissions. As a fast and reliable contact tracing is of foremost importance, several digital solutions have been proposed to match the need for personal information with privacy concerns [51, 52].

Other models have been applied to the emergence of COVID-19 in Belgium, either specifically [53, 54, 29] or in multi-country applications [55]. Using different model paradigms allows to focus on distinct aspects of the outbreak, like delay distributions of the clinical history of patients [29], a more detailed and age-specific handling of serological data with MCMC [54] or exploring individual-specific contact tracing options [53]. When evaluating intervention strategies with profound societal impact, ideally different models should be compared [56, 57].

In conclusion, we show the predicted impact of a phase-based relief of lockdown measures taken in Belgium. Through validation using empirical data on social contacts and the observed trajectory of the epidemic, our results suggest that the per-contact probability of infection has changed from pre- to post-lockdown. While economic and societal needs urge governments to relieve strict distancing measures and mobility restrictions, caution is required when evaluating different scenarios. Community contacts were found to be most influential, followed by professional contacts and school contacts, respectively, for an impending second wave of COVID-19. Regular re-assessment is crucial to adjust to evolving behavioral changes that can affect epidemic diffusion. In addition to social distancing, sufficient capacity for extensive testing and contact tracing is essential for successful mitigation.

## Data Availability

Demographic data is publicly available from Belgian Statistics. Surveillance data is publicly available and provided by the Belgian Scientific Institute for Public Health, Sciensano. Contact data are publicly available.

## Acknowledgements

This work received funding from the European Research Council (ERC) under the European Union’s Horizon 2020 research and innovation program (PC and NH, grant number 682540 – TransMID project, PL, NH, PB grant number 101003688 – EpiPose project). SA and NH gratefully acknowledge support from the Fonds voor Wetenschappelijk Onderzoek (FWO) (RESTORE project – G0G2920N). LW received funding from the Research Foundation Flanders (1234620N). The resources and services used in this work were provided by the VSC (Flemish Supercomputer Center), funded by the Research Foundation – Flanders (FWO) and the Flemish Government. We thank Giulia Pullano, Laura Di Domenico and Vittoria Colizza for useful discussions. The authors are also very grateful for access to the data from the Belgian Scientific Institute for Public Health, Sciensano.

## Author’s contribution

PC, PB and NH conceived the study. PC and PL contributed to the software development. PC, PL, SA, LW, PB and NH prepared the first draft of the manuscript. PC, SA, CF, OP, SAH and JW contributed to the data preparation and/or collection. All authors contributed to the final version of the paper and approved the final version of the manuscript.

Each member of the SIMID COVID-19 team contributed in processing, cleaning and interpretation of data, interpreting findings, contributed to the manuscript, and approved the work for publication.

